# Bibliometric Analysis of Manuscript Characteristics that Influence Citations: A Comparison of Six Major Family Medicine Journals

**DOI:** 10.1101/2020.04.13.20063719

**Authors:** Hamza Paracha, Amit Johal, Maida Tiwana, Dawood M Hafeez, Sabeena Jalal, Ateeq Rehman, Syed Asim Hussain, Faisal Khosa

## Abstract

**Objective:** The premise of our study was to investigate the characteristics of family medicine (FM) manuscripts that influence citation rate, capturing features of manuscript construction that are discrete from the study design.

**Design:** We conducted a cross-sectional study of published articles (n = 199), from January to June 2008, from 6 major FM journals with the highest impact factor. Annals of Family Medicine (IF = 1.864), British Journal of General Practice (1.104), Journal of American Board of Family Medicine (1.015), Family Practice (0.976), Family Medicine (0.936), and BMC Family Practice (0.815). Citation counts for these articles were retrieved using Web of Science filter on SCImago and 25 article characteristics were tabulated manually. We then predicted the citation rate by performing univariate analysis, spearman rank-order correlation, and multiple regression model on the collected variables.

**Results:** Using spearman rank-order correlation, we found the following variables to have significant positive correlation with citations: *number of references (r*_s_ and *p*-value, 0.31 and 0.001 respectively), *total words* (0.36, 0.001), *number of pages* (0.33, 0.001), *abstract word count* (0.17, 0.010) and *abstract character count* (0.16, 0.010). In a multivariate linear regression model: *number of references* (*p*-value = *0.010*, R^2^ = 0.06) *and multi-institutional* (*p*-value = *0.050*, R^2^ = 0.01) had a significant effect on citation rates.

**Conclusion:** Editors and authors of FM can enhance the impact of their journals and articles by utilizing this bibliometric study when assembling their manuscript.

## INTRODUCTION

There are about 2.5 million English and non-English scientific papers published per year.(Ware & Mabe, 2015) One measure of an article’s impact on the scientific community is the number of citations it receives.(Hirsch, 2005) The scientific rigor and significance of the article are important factors that determine its citation. However, scientific papers of significance may not attract the recognition they merit due to the assemblage of a manuscript. Furthermore, because citation count is contingent on discipline-specific trend and citation practices, the characteristics influencing it also differ between disciplines of medicine.(Moed et al., 1985) Hence, the purpose of our study was to determine various manuscript characteristics in Family Medicine (FM) and their impact on citation count. Our work has the potential to provide authors, journals, and institutions with knowledge on how to enhance their research’s visibility and improve the chances of its citation.

## MATERIALS AND METHODS

Our study aimed to complete a comprehensive analysis of variables that influence the number of citations.

### STUDYDESIGN

This cross-sectional study includes publicly available data and was exempt from institutional review board approval. We selected top 6 high impact factor (IF) FM journals with the most visibility and citations using the Web of Science filter on SCImago; Annals of Family Medicine (AFM) (IF = 1.864), British Journal of General Practice (BJGP) (1.104), Journal of American Board of Family Medicine (JABFM) (1.015), Family Practice (FP) (0.976), Family Medicine (FM) (0.936), and BMC Family Practice (BMC) (0.815). To ensure adequate time elapsed since publication and allow reliable citation reports, we derived the data from articles published in these journals between January 1, 2008, and June 30, 2008. A 6-month interval was selected to have ample time for a meaningful sample size. All original research and review articles were included while excluding case reports, editorials, commentaries, and letters to the editor. Our methodology has been validated in recent publications.(Hafeez et al., 2019),(Kim et al., 2020),(Shekhani et al., 2017) We have also updated the buildup of citation count on each article and our data analysis as of July 2020. As reported in a previous study by Lee et al., 90.7% of papers are not cited within the first year following publication, while the peak citation occurs 4 years after publication and then a gradual decline over 20 years.(Lee et al., 2005) Thus, the data selected from 2008 would allow enough time for a meaningful citation count to build up and provide substantial data for our analysis. We employed the Web of Science (Clarivate Analytics) to tabulate citation count for each article. The study variables of interest investigated for each article are shown in Table 1. Variables were selected carefully for analyses to consider the length of articles and abstract, types of study design, and its sample size as well as the accessibility of the journal.

### STATISTICAL ANALYSIS

Univariate linear regression against the number of citations was used to initially screen the variables. We calculated the p-values and identified the ones with *p*-values < 0.05. The variables that were considered statistically significant were then included in a multiple regression analysis. Multiple regression analysis was conducted for the variables that met the inclusion criteria. Those with 95% confidence interval (CI) and statistical significance (*p*-value <0.05) were noted. Positive interactions were assessed along with the presence of outliers.

We performed correlation analyses to better explain the association between our study variables and the number of citations. Dichotomous response variables were included in a point-biserial correlation analysis which is bivariate correlation analysis. Non-dichotomous variables were analyzed using Spearman’s rank-order correlation. A *p*-value of less than 0.05 was considered statistically significant.

For non-numerical values, ***study design***, and the ***continent of origin***, a non-parametric test, the Kruskal-Wallis test was used. Study designs were grouped as follows: 1. Cohort; 2. Surveys; 3. Case Series; 4. Randomized Control Trial (RCT); 5. Review; 6. Interventional; 7. Study design unspecified. The origin of articles was grouped based on continents: North America, Europe, Asia, and Australia.

## RESULTS

### STUDY CHARACTERISTICS

A total of 199 original and review articles that were published in the top 6 high-impact-factor (IF) journals were included in our study. Table 1 is a tabulated form of the characteristics of the studies included in our analysis. The important characteristics of the journals are in Table 2. The distribution of articles across the journals is available in Fig. 1A. The origins of the articles are based on the addresses of the corresponding authors are presented in Fig. 1B with its percentages from each continent. Study designs that were included in this analysis are based on the descriptions provided in each article and those that did not fall into any category were defined as “unspecified” in Fig. 1C. A full list of study variables, their medians, and interquartile ranges (IQR) is available in Table 3.

#### Citation Analysis

The median number of citations received per article was 20, with a minimum of 0 and a maximum of 225, (IQR: 28). Univariate linear regression analysis showed that citations of articles had an association with the variables ***abstract word count*** *(p=0.010)*, ***abstract character count*** *(p=0.010)****, number of pages*** *(p=0.001)*, ***number of references*** *(p=0.002)* ***and total words*** *(p=0.001)*. The variables that did not have a significant relation were excluded: ***number of words per title*** *(p=0.590)*, ***number of characters per title*** *(p=0.600)* ***number of authors*** *(p=0.800)*, ***number of tables*** *(p=0.310)*, ***number of figures*** *(p=0.160)*, ***sample size*** *(p=0.700)*. The variables that met the inclusion criteria were then used for multivariate regression analysis, which showed that the variable ***number of references*** *(p=0.010)* ***and multi-institutional*** *(p=0.050)* has a significant effect on citations.

For the dichotomous variables and their relation with the number of citations, we performed a point-biserial correlation and found that all the variables had negative association except one and that was for ***family medicine authors***, however, it lacked statistical significance. ***Multi-institutional*** studies showed a negative correlation but had a strong statistical significance (p=0.05), then the rest of the variables.

For our non-dichotomous variables, the test that we performed was Spearman’s rank-order correlation. We found that variables like ***abstract word count*** *(r*_s_ = 0.17 and p-value = *0.010)*, ***abstract character count*** *(0.16, 0.010)*, ***total words*** *(0.36, 0.001)*, ***number of references*** *(0.31, 0.001)* and ***number of pages*** *(0.33, 0.001)* had a *p-value (p<0.05)* and correlated with increased citations. Variables with no statistical significance were: **n*umber of tables*** *(0.03, 0.640)*, ***sample size*** *(0.02, 0.730)*, ***number of authors*** *(0.05, 0.410)*, and ***number of figures*** *(0.01, 0.840)*.

For variables like the ***continent of origin*** and ***study design***, we employed the Kruskal-Wallis test. This showed that articles from Europe had the highest citation, followed closely by North America. For the ***study design***, the highest citations were for the surveys, followed by RCT and cohort studies in our sample, but this has low significance as per our study and is not statistically significant. The studies that did not mention their study type were categorized as unspecified and those comprise almost 29 % of our sample. The highest numbers of citations per journal in descending order are AFM (2028), BMC (1091), BJGP (1056), FP (1036), JABFM (599), and FM (388). Medians are in the order of AFM, BJGP & JABFM followed by BMC and FM. The highest citations were observed by AFM; however, the citations were more significantly distributed in the BMC, BJGP, and FP.

## DISCUSSION

According to our study results, characteristics relating to the length of the article, such as total word count, number of pages, and references, showed a statistically significant positive correlation with an increased number of citations. Antoniou et al. also reported a positive correlation between the length of the article, number of references, and the inclusion of study design in the title with increased citation count in their analysis, even after adjusting for several potentially confounding variables.(Antoniou et al., 2015) We found that the total words and the number of pages within a dermatology manuscript had the strongest positive correlation for a higher citation count.(Kim et al., 2020) The results of this study can benefit authors who may improve the citation of their articles by utilizing this bibliometric study when assembling their manuscript. This may signify that longer articles may perhaps contain more ideas, information, and greater diversity of results attracting opportunity for higher citation rates.

Similarly, the number of figures and the number of tables showed a positive correlation only. While these variables also pertain to the length of the article; in our study, their statistical significance was not observed. Shekhani et al. suggested that having a greater number of contributors or authors on a radiology scientific article may increase its quality, length, references, and citations.(Shekhani et al., 2017) However, our data for FM journal articles only correlates to the length and references of the article.

The abstract word count and abstract character count had a positive correlation with statistical significance, while structured abstract displayed a negative correlation with no statistical significance. Previous studies have shown that the format of the abstracts is influenced by the publishing journal, the instructions for authors and processes of review and editing.(Nakayama et al., 2005) The instructions prescribed by the journal are essential to maintain the quality of the abstracts. Abstracts provide an overview and introduce the reader to the ideas presented in the manuscript. Allowing authors to have an elongated abstract would provide them the freedom to freely describe their study findings. Editors may also benefit their journal and authors by supporting and promoting the use of longer abstracts.

Having FM articles published with open access had a negative correlation with no statistical significance. Davis PM reported that, although open access publishing may reach more readers, there was no evidence found to support a citation advantage for open access articles cited within the first year after publication, when compared with subscription-access control articles cited within the first 3 years of publication.(Davis et al., 2008) The data collected for our study was from articles published more than 10 years ago. Although, journal articles with open access may be readily available and accessible, which could potentially allow greater visibility and audience, therefore, higher chances for citations, our study did not show such results.(Davis, 2011)

Aksnes and Falagas et al. reported that, generally, team-authored studies and multi-institutional collaborations received increased citations.(Aksnes, 2003),(Falagas et al., 2013) Our study showed a slight positive correlation with no statistical significance of higher citation count with an increased number of authors, yet, there was a statistically significant negative correlation with multi-institutional studies. Some articles pertaining to FM may be published in different subspecialty journals. Although correlation does not equal causation, we cannot hypothesize the reason behind the strong negative correlation to multi-institutional collaboration with citation count.

Words and characters per title also had a positive correlation with no statistical significance contrary to a prior publication in psychiatry.(Hafeez et al., 2019) These results also differ from another study by Jacques et al. that stated the length of the title, the presence of a colon in the title, and the presence of an acronym showed a positive correlation.(Jacques & Sebire, 2010) Our study displays a negative correlation with no statistical significance for punctuation in the title which also contrasted Jacques et al.(Jacques & Sebire, 2010) Other studies have shown that shorter titles may potentially be easier to understand and results in an increased citation.(Paiva et al., 2012),(Habibzadeh & Yadollahie, 2010) Some authors maintain that many of the literature searches are performed electronically on sites such as PubMed or Google. Naturally, a longer title can lead to an increased chance of the article appearing in a search result and therefore can increase the chances of a researcher finding the work and citing it. On the other hand, misleading titles may skew results in the other way.(Jacques & Sebire, 2010) (Chokshi et al., n.d.)

### Limitations

Our study is subject to selection bias as the six journals selected were those with high-impact factor. Results may not be representative of all FM journal articles. The citation count for the first ranking journal is nearly double compared to each of the following three journals. This has the potential to skew the results, which affects correlation in smaller sample sizes than larger ones.(Goodwin & Leech, 2006) Our sample size, n=199 from six journals, and the time frame of six months may seem appropriate but the inclusion of more journals, larger sample size, and a longer time frame could potentially yield more powerful results.

Authors cite papers for a multitude of reasons ranging from supporting their study findings and text, referring to a particular methodology, and to discuss examples of deceptive results or faulty methods.(Belter, 2015) These citations represent the usefulness of the article to other scientific authors.(Belter, 2015) However, bibliometric indicators count all citations equally, regardless of the reasons and cannot account for the intention behind the citation.(Belter, 2015) This may present as a potential limitation in using bibliometric indicators to influence citation rates.

The statistical significance of the data does not inevitably interpret a correlation being the representative of causation. Our results that displayed statistically significant results had very weak correlation coefficients. If we had included more journals in our study, it is possible that due to increased variability in the data and lesser effect of outliers, our results may have a stronger correlation between manuscript characteristics and citations. The difference in the median number of citations between the first ranking journal and the following three journals is almost twice; and the 5th ranking journal to be approximately 4 times, there is a potential for skewed data results. According to Goodwin et al, the effect of skewed data on correlation becomes larger in a smaller dataset than in a larger one.(Goodwin & Leech, 2006) Moreover, a stronger correlation with citations would be better explained by scientometric analysis, which measures and analyzes science itself. Naveed et al. reported that certain topics attract more interest in a specific time period while analyzing citation trends over four decades. Factors that may influence these citation trends are time period, area of research, and the topics being studied are better analyzed through scientometric analysis.(Waqas et al., 2017) There is potential for exploring citation correlation with multi-institutional collaborates over an extended period of time to better explain the trends over time.

Human error must be addressed due to the manual collection of data for each article. Databases such as Web of Science (WOS), Scopus (Elsevier), and Google Scholar (Google) also differ in citation counts. We chose WOS articles through SCImago as our database because it offers a strict evaluation process. It also provided the most reliable and integrated citation metrics from multiple sources in a single interface.(Falagas et al., 2008) Our research is limited in the context that FM research articles are scattered among FM journals and even more extensively through non-FM journals making it difficult to fully incorporate all articles with vast numbers of citations.(Dunikowski & Freeman, 2016)

## CONCLUSION

Based on the result of our bibliometric analysis, authors of articles publishing in FM journals can improve their research visibility and citation count by paying close attention to certain article characteristics. Our study findings suggested having higher total words, number of pages, references, abstract word count, and abstract character count showed a statistical significance. The results also showed a negative correlation with statistical significance in the manuscript assembled with multi-institutional authors. Our results suggest that bibliometric knowledge is beneficial for authors and journals to know which manuscript characteristic may or may not help to enhance the visibility and citation count of their articles.

## Data Availability

Data is available upon request.

## Funding Statement/Disclosures

Dr. Khosa is the recipient of the AFMC-May Cohen Equity, Diversity and Gender Award (2020); Canadian Association of Radiologists – Young Investigator Award (2019); Rising Star Exchange Scholarship Program of French Society of Radiology (2019) and Humanitarian Award of Association of Physicians of Pakistani Descent of North America (2019). The authors have no relevant disclosures.

## Notes

### Competing Interest Statement

The authors have declared no competing interest.

### Funding Statement

No external funding was received.

